# Reliability of remote self-administered web-based digital cognitive measures and comparison to in-person neuropsychological tests: Stricker Learning Span, Symbols Test and the Mayo Test Drive Screening Battery Composite

**DOI:** 10.1101/2025.09.26.25336467

**Authors:** Morgan A. Hughes, Ryan D. Frank, Rita L. Taylor, Winnie Z. Fan, Teresa J. Christianson, Walter K. Kremers, John L. Stricker, Mary M. Machulda, Jason Hassenstab, Michelle M. Mielke, John A. Lucas, Paula A. Aduen, Gregory S. Day, Neill R. Graff-Radford, Clifford R. Jack, Jonathan Graff-Radford, Ronald C. Petersen, Nikki H. Stricker

## Abstract

**INTRODUCTION:** We describe the reliability of remote self-administered digital cognitive measures completed via the Mayo Test Drive (MTD) web-based platform.

**METHODS:** 1,846 participants (mean age=70, SD=12, range 31-101; 48% male; 96% White; 99% non-Hispanic; 97% cognitively unimpaired) with 2-4 complete MTD sessions at ~7.5-month intervals were included. Test-retest reliability was assessed using single-rating, absolute-agreement, and two-way mixed intraclass correlation coefficients (ICCs) with 95% confidence intervals. ICCs for in-person-administered traditional neuropsychological measures were compared to MTD for a subset of 244 participants.

**RESULTS:** Reliability was good for the MTD Composite [total ICC = 0.79 (0.77, 0.80)], and moderate-to-good for the primary outcome variables for each MTD subtest [total ICCs 0.70-0.83 for Stricker Learning Span and Symbols]. The reliability of the remote self-administered MTD was similar to in-person-administered cognitive measures.

**DISCUSSION:** MTD showed moderate-to-good reliability, supporting its use in longitudinal monitoring.

## Introduction

Mayo Test Development through Rapid Iteration, Validation and Expansion (DRIVE, MTD), is a web-based, multi-device (i.e. smartphone, tablet, and computers) digital platform used for remote self-administration of cognitive assessments with high usability.^1^ The MTD cognitive screening battery includes the Stricker Learning Span (SLS) and Symbols (SYM) subtests, and is typically completed in 15-20 minutes.^1^ The SLS is a computer adaptive word list memory test and the SYM test measures processing speed and working memory. The MTD screening battery composite score (MTD_Comp_) is a combination of the SLS and SYM test that shows robust associations with the Mayo Preclinical Alzheimer’s disease Cognitive Composite (Mayo-PACC)^2^ and multiple neuroimaging biomarkers including amyloid PET, tau PET, hippocampal volume, and white matter hyperintensity volume.^3^ Normative data for remote self-administration of MTD are available.^4^

The validity of MTD and its subtests are well-supported by prior publications.^3–5^ Reliability is an additional key consideration for remote digital cognitive assessments.^6^ Test-retest reliability refers to the ability to replicate a test over time, or how consistent test results are within an individual across sessions.^7^ Because MTD subtests use a randomized item selection approach, test-retest reliability for MTD subtests also includes alternate-form reliability.^5^ Remote assessments can be completed in multiple environments, which may lead to a higher likelihood of distractions or interruptions that may increase variability in performance and compromise reliability. Reliable remote self-administered digital cognitive assessments would expand options for monitoring cognition over time.

The aim of the current study was to characterize the reliability of MTD. We compared the reliability of remotely administered MTD measures to traditional in-person-administered neuropsychological measures. We also examined the impact of demographics (age, education, sex), device type / input source, subtest interference^1^, and location on reliability.

## Method

### Participants and Study Procedures

MTD study participants were primarily recruited from the Mayo Clinic Study of Aging (MCSA), which is a prospective, population-based study of aging within Olmsted County, MN.^1,4^ MCSA study visits occurred every 15 months (or every 30 months for individuals <50 years of age) and included a physician exam, completion of the Clinical Dementia Rating® (CDR),^8^ and in-person neuropsychological testing.^9^ Clinical diagnoses of cognitively unimpaired (CU), mild cognitive impairment (MCI), or dementia were determined by consensus agreement, referencing published diagnostic criteria.^10,11^ MCSA participants were invited to complete an MTD session the week following their MCSA study visit and every 7.5 months between study visits.^1^ See Supplemental Online Resources for additional time interval details. Some participants were also recruited from the Mayo Alzheimer’s Disease Research Center, as previously described.^1,4^

This study was conducted in accordance with the Declaration of Helsinki. The MTD study was approved by the Mayo Clinic IRB. Oral consent (that includes review of consent elements via email) was obtained.^1,4^

### MTD Sessions

MTD subtests and variable derivation have been previously described in detail.^3,4,12,13^ The SLS includes five adaptive learning trials and a delayed recognition memory trial.^5^ During learning trials, participants are visually presented with one word at a time and asked to identify the presented words in a 4-word recognition format at the end of each trial. The delay trial is presented after the SYM test. SLS Sum of Trials (SLS_Sum_) is the primary outcome variable (trials 1-5 total correct + delay correct).

The SYM test requires rapid matching of symbol pairs. There are 12 items in each trial, and 4 trials are completed sequentially.^3^ MTD SYM is an adaptation from the Symbols test administered in the Ambulatory Research in Cognition (ARC) App^14^; the primary outcome variable is average correct item response time in seconds (SYM_RT_). We provide an additional primary MTD SYM variable that weights SYM_RT_ by accuracy, the SYM accuracy-weighted score (SYM_AW_). MTD_Comp_ is the sum of SLS_Sum_ and SYM_AW_.^3,4^

Secondary variables (see Table 2) can be considered for use as outcome variables for research studies and for clinical interpretation as needed (e.g., when immediate learning and delayed memory performance differentiation is emphasized). We included an alternative, z-score based version of the MTD_Comp_ (MTD Composite-z) that has been previously described.^3^ This may be used in research as an alternative outcome variable but no normative data are available for this version of the composite because it is derived from z-scores and thus is sample specific.

Secondary process variables (see Table 2) are expected to have lower reliability because they represent single trial variables or are expected to have non-normal distributions.^15^ Secondary process variables should not be used as outcome variables for research studies, but are available to characterize patterns of performance, an individual’s approach to testing (i.e., “process”), or to allow alternative metrics if an interruption impacts one or more primary variables (e.g., if an individual is interrupted during one or more SLS or SYM trial).

### Inclusion Criteria for Primary Analyses

Inclusion criteria for the primary analyses were: 1) a complete baseline MTD session (i.e. MTD_Comp_ not missing) and at least one complete and consecutive follow-up session; 2) a valid session, as previously defined^1^ or per data cleaning/session review; 3) absence of dementia at baseline. MTD sessions completed between 5/25/21 and 6/15/25 were included.

### Sample Selection for Comparison to In-Person Traditional Neuropsychological Tests

To compare the reliability of traditional in-person-administered neuropsychological tests to self-administered MTD measures, the sample was limited to participants newly enrolled into the parent study to ensure participants were naïve to both MTD and in-person neuropsychological tests. Because in-person-administered neuropsychological testing occurred approximately every 15 months, the 2^nd^ in-person visit occurred around the same time as the 3^rd^ MTD session (MTD is completed every 7.5 months). Comparisons to in-person testing focused on the first two MTD sessions (sessions 1-2) for consistency of the number of administrations between remote and person-administered measures, but session 1-3, 2-3, and total reliability across all 3 sessions are also reported to show a similar time interval. Comparisons focused on the following in-person-administered measures: (1) the Kokmen Short Test of Mental Status (STMS)^16^, which is a multi-domain global cognitive screening measure (similar to the Mini Mental State Examination), and (2) the Mayo-PACC, which is an average of z-scores from the Rey’s Auditory Verbal Learning Test (AVLT) sum of trials, Trail Making Test B (inversed) and animal fluency.^2^ We additionally compared select subtest measures from Mayo-PACC considered most similar to MTD subtest measures (AVLT sum of trials and SLS_Sum_; SYM_AW_/SYM_RT_ and Trails B).

### Sample Selection for Subgroup Analyses

To examine potential MTD reliability differences by demographics and by factors relevant for remote assessment (device type, subtest interference, and location), the primary analytic sample was limited to CU participants before creating subgroups. Dichotomous groups were created to facilitate comparisons for the following: age (age <70-vs ≥70-years old), sex, education (<16 vs ≥16 years of education), device and input source consistency (participants who consistently used the same device/input source for all sessions vs. participants who switched types across sessions), potential test interference (no endorsed potential interference during any session vs ≥1 potential distraction or interference during a session)^1^, and location (consistently took all MTD sessions at home vs. location varied across sessions).

### Statistical Methods

Patient demographics were summarized using counts and percentages for categorical variables and means and standard deviations for continuous variables. Comparison of data distributions across groups were performed using chi-square tests for categorical variables and 2-sample t-tests for continuous variables. The ICCs were calculated as 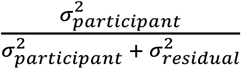 from a random effects model with random effects participant intercepts (unadjusted ICCs) and fixed effects for age at assessment, sex, and education (adjusted ICCs). Unadjusted ICCs are typically reported in studies reporting reliability of cognitive test data but are sample-specific. We therefore also provide adjusted ICCs. Bootstrap methods using 1000 simulations were used to estimate the standard error used in the confidence limits for ICCs. We also derived Pearson correlation coefficients and their 95% CI. Raw scores were Winsorized at the 1^st^ and 99^th^ percentile to lessen the influence of potential outliers. We applied reliability descriptives as defined by Koo et al.^7^ (ICCs < 0.5 = poor; ICCs 0.5-0.75 = moderate; ICCs 0.75-0.90 = good; and ICCs > .90 = excellent). Unadjusted and adjusted ICCs (where the adjustment terms were included as fixed effects) were calculated in independent subsets of the overall sample for subgroup analyses (e.g. males vs females) and compared using the properties of variance distributions (i.e. Var(X-Y) = Var(X) + Var (Y) – Cox(X,Y) = Var(X) + Var(Y) in independent samples) and bootstrap methods; results interpretation focused on adjusted *p*-value comparisons for subgroup analyses (adjusting for demographic variables). Analyses were conducted using R version 4.4.1. Statistical tests were 2-sided; p-value <0.05 was considered statistically significant.

## Results

### Participant Demographics

1,846 participants met inclusion criteria and were included in the primary analyses. Participants were 31-101-years-old at baseline and completed 2-4 MTD sessions (Table 1). Time between sessions was 7-9 months. Nearly all (99.5%) MTD sessions were completed remotely using the participants’ personal devices (61.9% computer, 22.6% smartphone, 15.2% tablet, 0.3% unknown/other). Most participants were White (95.9%), non-Hispanic (99.0%), college educated (mean years of education=15.7, SD=2.3, range 6-20), older adults (mean=69.7, SD=12.1) and CU (96.7% CU; 3.3% were diagnosed with MCI).

**Table 1.**
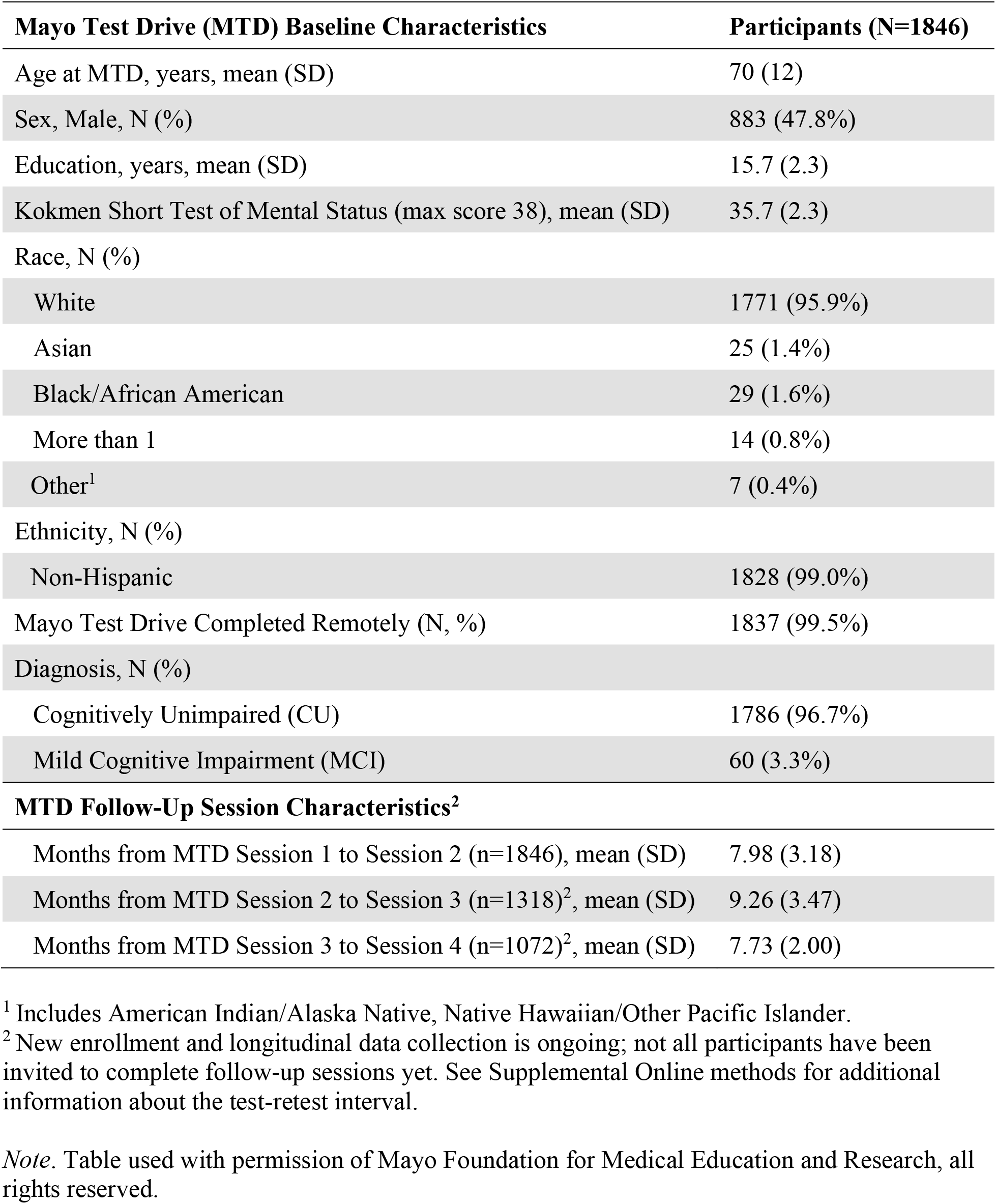
Demographic and other characteristics of the sample.

### MTD Reliability

Reliability was good for MTD_Comp_ (total ICC = 0.79; see Table 2 for session-to-session ICCs and 95% CIs), and moderate-to-good for primary outcome variables for SLS and SYM (total ICC: SLS_Sum_ = 0.75, SYM_AW_ = 0.70, SYM_RT_ = 0.83; see Figure 1). Weighting by accuracy reduced the reliability of the SYM test and MTD_Comp_; SYM_RT_ had a higher total ICC (0.83) than SYM_AW_ (0.70). Similarly, the alternative MTD Composite-z (that includes the SYM_RT_ instead of SYM_AW_) showed subtly higher ICC values relative to MTD_Comp_ with total ICCs of 0.81 and 0.79, respectively. SLS sum of trials had subtly higher reliability (0.75) than separate secondary measures of learning (1-5 total correct = 0.73, max span = 0.70) or delay (0.72). Alternative time-based SYM variables showed nearly identical reliability values (e.g., total ICC = 0.83 for SYM_RT_ and average seconds across all four trials, and total ICC = 0.82 for middle two trials average seconds), and averaging across SYM trials yielded higher ICCs compared to single trial ICCs. All secondary variables had moderate-to-good reliability coefficients. As expected, some secondary process variables based on a single trial or with restricted ranges and non-normal distributions had poor reliability coefficients (SLS Percent Retention, SYM Accuracy/Total Correct, SLS Trial 1, SLS Trial 2), but most secondary process variables had moderate-to-good reliability coefficients. Pearson correlations were highly consistent with ICC reliability values; unadjusted ICCs of all primary variables were within .02 of the Pearson correlation coefficients (see Figure 1 and Supplemental Table 2).

**Table 2.**
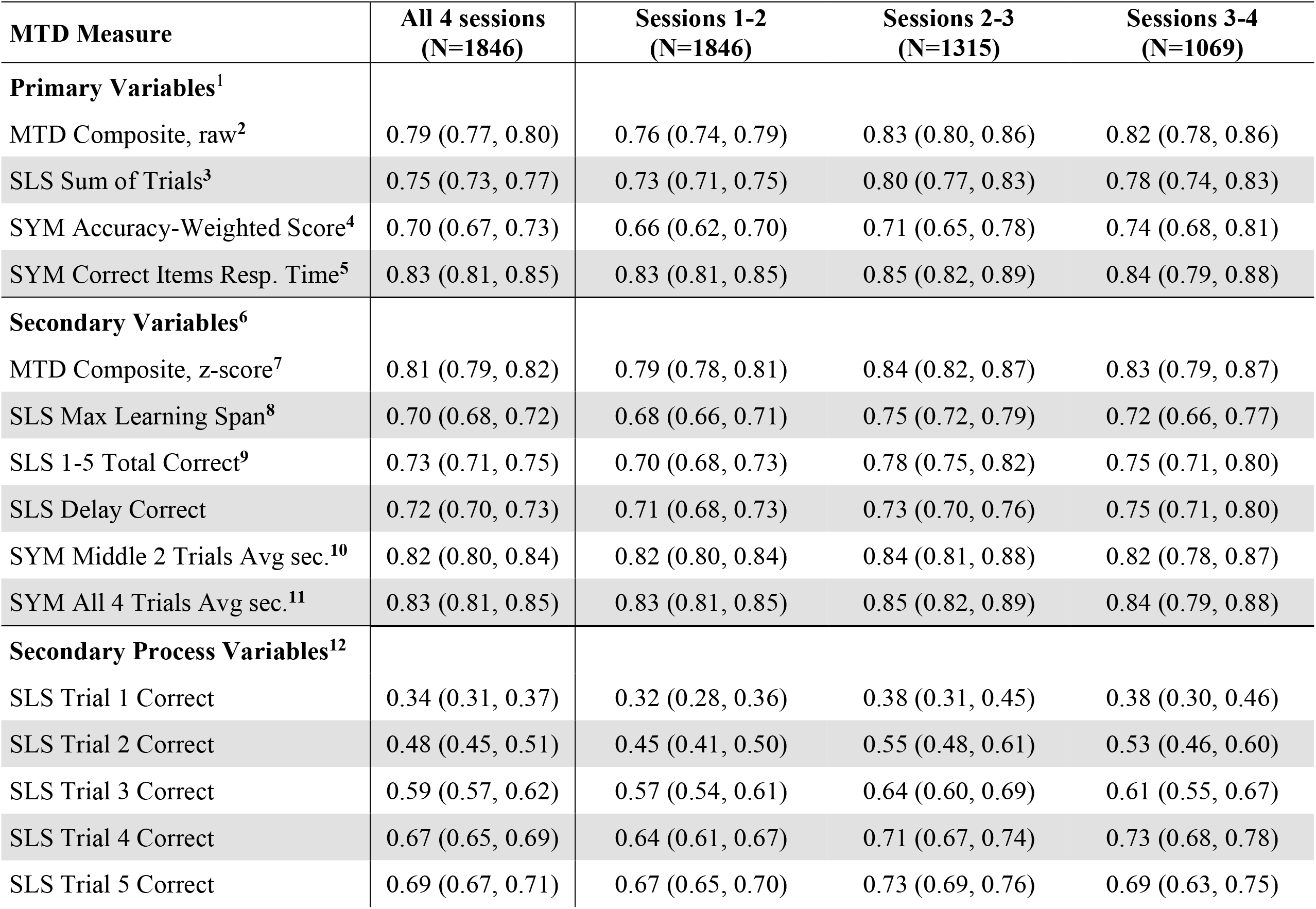
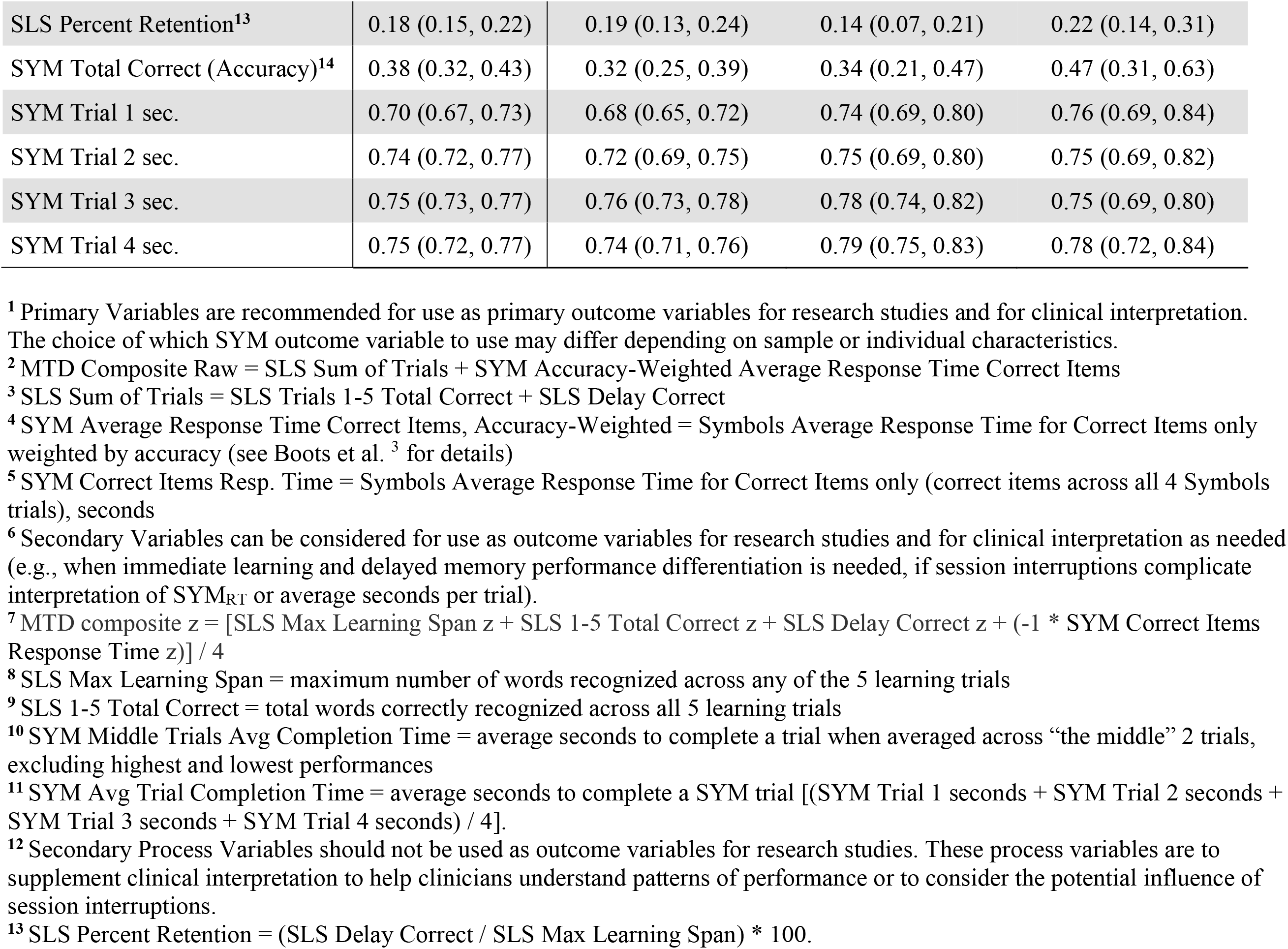
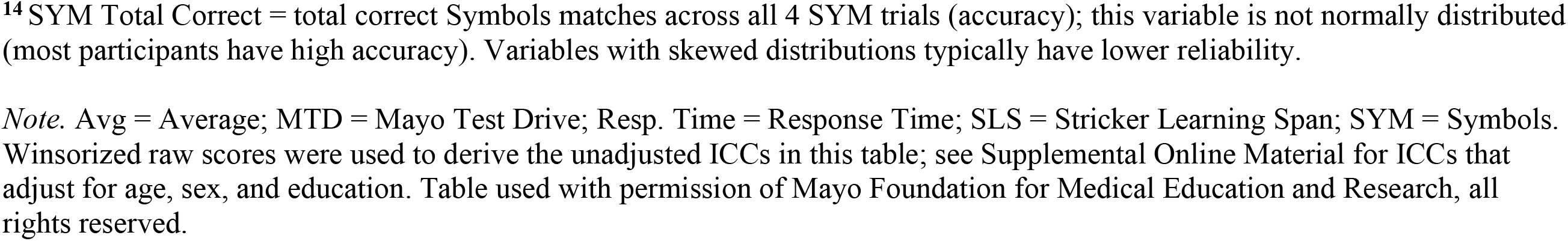
Mayo Test Drive Intraclass Correlation Coefficients (ICC, 95% Confidence Interval) for all participants averaging across all sessions and for adjacent single test-retest session pairs.

**Figure 1.**
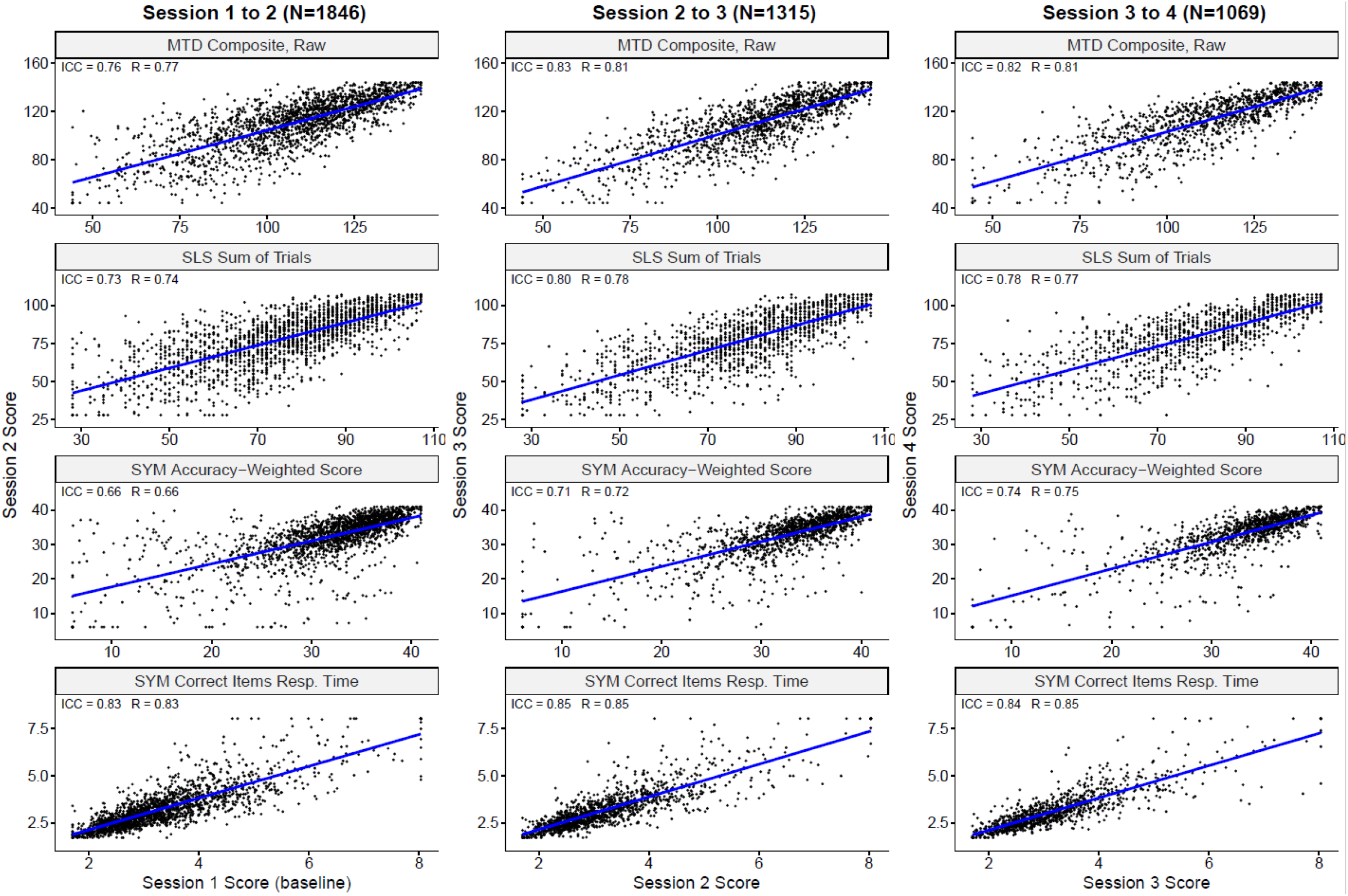
Scatterplots showing single session test-retest reliability for primary MTD variables for adjacent test sessions in all participants. ***Note***. Unadjusted intraclass correlation coefficients and Pearson correlation coefficients for Winsorized raw scores are reported for each scatterplot (see Tables 2 and S2 for 95% confidence intervals, and Table S3 for Pearson correlation coefficients using non-Winsorized scores). MTD = Mayo Test Drive; Resp. Time = Response Time; SLS = Stricker Learning Span; SYM = Symbols. Figure used with permission of Mayo Foundation for Medical Education and Research, all rights reserved.

### MTD Reliability Compared to Reliability of In-Person Administered Tests

Table 3 shows ICC values for in-person and remote MTD testing. When comparing two traditional in-person sessions (baseline to ~15 months) and two MTD sessions (baseline to ~7.5 months), MTD_Comp_ ICC was similar to that of STMS and Mayo-PACC (*p*’s > .05; Supplemental Table 4). Subtest comparisons showed that SLS sum of trials ICC was similar to AVLT sum of trials ICC (*p* = .15), SYM_AW_ ICC was lower than Trails B ICC (*p* = .04), and SYM_RT_ ICC was higher than Trails B ICC at trend level (*p* = .051). See Supplemental Table 5 for alternative comparisons (e.g., comparing 2 in-person sessions to the ICC across all 3 MTD sessions or to MTD sessions 1 and 3 to capture the same time interval). Across all comparisons, there is a trend for reliability of MTD composite to be higher than the STMS, and there is a trend for reliability of Trails B to be higher than SYM_AW_ (but similar to SYM_RT_).

**Table 3.** Intraclass Correlation Coefficients (ICC, 95% Confidence Interval) for remote self-administered Mayo Test Drive (MTD) and in-person-administered traditional neuropsychological measure reliability for a subset of 244 participants newly enrolled to the parent study.

| **In-Person-Administered Traditional Neuropsychological Measures** | **Visits 1-2<sup>1</sup>** |  |  |
| --- | --- | --- | --- |
| Kokmen Short Test of Mental Status<sup>2</sup> | 0.67 (0.58, 0.75) | - | - |
| Mayo-PACC<sup>3</sup> | 0.80 (0.75, 0.85) | - | - |
| AVLT Sum of Trials<sup>4</sup> | 0.78 (0.73, 0.83) | - | - |
| Trails B | 0.74 (0.66, 0.82) | - | - |
| Animal Fluency | 0.62 (0.54, 0.70) | - | - |
| **Remote Mayo Test Drive Measures** | **Sessions 1-2** | **Sessions 2-3** | **Total ICC**<br>**All 3 Sessions** |
| MTD Composite, raw | 0.75 (0.70, 0.81) | 0.78 (0.73, 0.83) | 0.76 (0.72, 0.81) |
| SLS Sum of Trials | 0.72 (0.66, 0.79) | 0.76 (0.70, 0.82) | 0.74 (0.69, 0.79) |
| SYM Accuracy-Weighted Score | 0.61 (0.50, 0.71) | 0.59 (0.47, 0.71) | 0.60 (0.50, 0.70) |
| SYM Correct Items Response Time | 0.83 (0.78, 0.88) | 0.78 (0.72, 0.84) | 0.80 (0.75, 0.85) |

<sup>1</sup> Months between visits for in-person measures was 16.0 (SD=1.7). The second in-person visit occurred around the time of the 3<sup>rd</sup> MTD session.
<sup>2</sup> Kokmen Short Test of Mental Status is a multi-domain screening measure similar to the MMSE
<sup>3</sup> Mayo-PACC = Mayo Preclinical Alzheimer's disease Cognitive Composite (average z of Auditory Verbal Learning Test sum of trials, animal fluency, and inversed Trails B)
<sup>4</sup> AVLT Sum of Trials = Rey's Auditory Verbal Learning Test trials 1-5 correct + trial 6 correct + 30-minute delayed recall correct
*Note.* Winsorized raw scores were used to derive the unadjusted ICCs in this table; see Supplemental Online Material for ICCs that adjust for age, sex, and education, demographic characteristics of this subsample, and for *p*-values for direct ICC comparisons (in-person vs. remote measures). Table used with permission of Mayo Foundation for Medical Education and Research, all rights reserved.

### Reliability by Demographic Variables and by Factors Relevant to Remote Self-Administered Assessments

The sample was limited to CU participants (N=1,786) prior to subgroup comparisons. See Supplemental Table 5 for subgroup ICC (95% CI) values, unadjusted *p*-values, *p*-values adjusted for demographic variables (*p*_adj_), and subgroup characteristics for these analyses. Reliability scores were similar across MTD primary variables when comparing education (<16 years education vs. ≥16 years education; *p*_adj_’s ≥ 0.35) or sex (males vs. females; *p*_adj_’s ≥ 0.30) subgroups.

Most participants (73%, n=1310) used the same device/input source combination for all MTD sessions, typically a computer and mouse (61.9%). There were no significant differences between reliability scores for consistent versus inconsistent device/input source groups for most MTD primary variables (*p*_adj_’s ≥ 0.13). However, SYM_RT_ showed higher reliability for the consistent group relative to the inconsistent device-input source group (*p*_adj_ = 0.049)

597 CU participants endorsed potential interference during at least 1 testing session (interference group). There were no significant reliability differences between the interference^1^ and no interference groups (*p*_adj_’s > 0.10).

Most participants (85%, n=1516) consistently completed all MTD sessions at home. MTD_Comp_ and SLS_Sum_ showed no reliability differences between consistent and inconsistent location groups (*p*_adj_’s > 0.14); however, both SYM primary variables showed higher reliability coefficients when all sessions were completed at home (*p*_adj_ < 0.01 for both).

No significant effect of age on reliability was observed for SLS_Sum_ and SYM_RT_ (*p*_adj_’s > 0.05). However, the ≥70 age group showed higher ICC compared to the <70 group for MTD_Comp_ and SYM_AW_ (*p*_adj_’s < 0.03). Chi-square analysis showed the ≥70 group was more likely to take all MTD tests at home compared to the <70 group (*p* < 0.001), thus greater location consistency of the 70+ group is suspected to drive this age difference.

## Discussion

This study provides a detailed characterization of the test-retest reliability of the remote, self-administered digital MTD screening battery composite and its subtests with 7-9-month test-retest intervals across 2-4 test sessions in a large community-based research sample. MTD_Comp_ had good reliability overall. Primary and secondary subtest outcome variables had moderate-to-good reliability, similar to our prior report of 2-week test-retest reliability of SLS and SYM subtest variables in an initial pilot study.^13^

MTD has favorable reliability relative to other digital cognitive assessments that use a single session approach for a given timepoint, including those administered in clinic and remotely. For example, Stricker et al.^17^ examined the reliability of the in-clinic self-administered Cogstate Brief Battery with a 7.5-month test-retest interval in CU individuals and showed poor-to-moderate reliability (ICCs from 0.36-0.59) for a learning/memory measure (one card learning (OCL) accuracy). Kochan et al.^18^ reported poor at-home self-administered 1-month test-retest reliability for Cogstate OCL (ICC = 0.43) and Cambridge Brain Sciences Paired Associates (ICC = 0.30), though use of various cross-battery global composites yielded good reliability (ICC’s 0.82-0.85). Feenstra et al.^19^ reported moderate reliability for word list learning (ICC = 0.59), delayed recall (ICC = 0.50) and recognition (ICC = 0.70), moderate reliability for a measure similar to TMT (Connect the Dots ICCs = 0.67-0.71), and good reliability (0.83) for a 7-subtest composite using the personal-computer-based self-administered Amsterdam Cognition Scan 1-hour battery in 248 healthy adults completed twice within 6 weeks at home. Rigby et al.^20^ found similar reliability to MTD subtest results in their cohort of CU and MCI participants when using the in-person-administered National Institute of Health Toolbox-Cognition Battery (NIHTB-CB) in a clinic setting twice within four months. The NIHTB-CB picture sequence memory test (ICC = 0.65) and the pattern comparison processing speed (ICC = 0.85) showed moderate-to-good reliability, and the Fluid Composite showed good reliability (ICC = 0.85).

Reliability increases as the number of items, trials, tests, and assessments increases.^2,21^ Thus, “one-shot” single session approaches tend to have lower reliability relative to paradigms that use the average across several very short sessions completed several times a day for several days or weeks (i.e., ecological momentary assessment)^14^, or longer sessions repeated once daily for several days.^22,23^ Although individual sessions within these multi-test approaches are brief, cumulatively, they represent more testing time for a given timepoint compared to brief one-shot assessments like MTD. For example, one 15-minute daily session for 6 days represents 90 minutes of testing^22^ and 4 daily 30-60 second sessions over 7 days represents 42-84 minutes of testing^14^. Nicosia et al.^14^ found excellent reliability when assessing the ARC app in a cohort including CU and very mildly impaired (global CDR 0.5) participants After testing in cycles of 4x a day for 7 consecutive days, the ICCs of each ARC subtest was >0.85 and the reliability of the ARC composite scores were excellent (>0.90) between the initial testing cycle and both a 6-month and a 1-year follow-up cycle. Nicosia et al.^14^ also showed that SYM_RT_ reliability reaches 0.88 when averaging across 3 sessions; these data informed the decision to include 4 consecutive SYM trials within a single MTD session as an adaptation of ARC Symbols for inclusion in a “one-shot” assessment paradigm. The current study similarly demonstrates that averaging across ≥2 SYM trials yields ICC values >0.8, whereas single-trial SYM reliabilities ranged from 0.69-0.76. Weighting by accuracy reduced the reliability of SYM, but it helped capture non-speed related aspects of SYM test performance and facilitated creation of the MTD_Comp_.^3^

Reliability of the remote self-administered MTD_Comp_ was similar to an in-person-administered mental status screening measure (STMS) and a composite from traditional in-person-administered neuropsychological tests (Mayo-PACC). Subtest-level comparisons showed that the SLS had similar reliability compared to the AVLT, SYM_AW_ had lower reliability relative to Trails B, and SYM_AW_ had similar reliability relative to Trails B. Direct comparisons to in-person tests focused on comparing test 1 to test 2 ICCs to allow comparison on the same number of assessments, though this resulted in differing time intervals (7.5 months for MTD vs. 15 months for in-person measures), which is an important limitation of these findings. Results were broadly similar when comparing in-person-administered tests to total ICC that considers all 3 timepoints and to comparisons using only MTD sessions 1 and 3 to achieve a similar follow-up interval, though across comparisons there was a trend for reliability of MTD composite to be higher than the STMS. The reliability of the STMS in the current study is comparable to reliability coefficients reported for other global mental status screening measures. For example, Pedraza et al.^24^ reported DRS reliability ranging from 0.54-0.71 (Pearson correlation coefficients for 16-24- and 9-15-month intervals, respectively) in CU older adults. MMSE reliability is consistently reported to be poor, including in 119 CU older adults over a test-retest interval of 1.6 years (Spearman correlation coefficient = 0.31)^25^, and in 65 CU participants over a test-retest interval of 83 days (Spearman correlation coefficient = 0.35).^26^ To our knowledge, no other group has compared the reliability of remote and in-person-administered traditional cognitive measures in the same participants, making this a strength of the current study.

Remote digital assessments offer several advantages to patients, including increased cost-effectiveness, time efficiency and convenience. In addition, remote digital assessment may reduce the “white-coat effect” that may occur with in-person-assessments^28^ and allow measures to be easily repeated over time.^18,19^ The potential benefits of remote digital assessment compared to person-administered measures such as the Mayo-PACC or STMS is demonstrated by a post-hoc power analysis using the results of this study. Because MTD can be completed more frequently remotely, a smaller number of patients is needed to detect effects in clinical trials. We used data derived from MTD vs in-person-administered measure comparisons to calculate the estimated magnitude of this difference. For example, if using the same visit/session frequency as used in the current study, for a clinical trial of 5 years there would be 5 in-person testing sessions (baseline, 15 months, 30 months, 45 months, end of study), whereas MTD would have 9 remote sessions (every 7.5 months). To obtain 80% power to detect a slope difference of 0.10 standard deviations, MTD would require 328 participants, Mayo-PACC would require 380 participants, and Kokmen STMS would require 418 participants, assuming a type I error rate of 0.05 and a ratio of subject-specific random slope variation to be 0.1 from a random effects model with random participant slopes and intercepts. Thus, MTD would reduce the sample size needed by 13.7% (Mayo-PACC) to 21.5% (STMS).

Although remote digital assessments have many benefits, disadvantages are recognized. First, a lack of environmental control may lead to inconsistency and inaccurate measurements. Encouragingly, our results show that the reliability of MTD is minimally affected by self-reported potential test interferences. Second, some older adults may not have access to certain devices or may be uncomfortable using technology by themselves.^1,29^ Multi-device platforms may help lessen this concern by allowing use of any available device the patient or participant are most comfortable with. The current results show that the reliability of the MTD is minimally affected by variation in device/input source type across sessions, although device consistency is recommended when feasible and location consistency may be important. It is important to note that the current study’s participants were predominately CU, White, non-Hispanic, and highly educated. Further research is needed to ensure generalizability of MTD to racial, ethnic, and socioeconomically diverse populations.

In summary, the current study demonstrates that the MTD screening battery is a reliable assessment for tracking cognition over time, with moderate-to-good reliability. MTD reliability is comparable to in-person-administered neuropsychological tests as well as other digital cognitive assessments and is favorable compared to in-person mental status screening measures. Future studies are needed to determine the sensitivity of MTD to detect longitudinal cognitive decline.

## Funding & Acknowledgements

Research reported in this publication was supported by the National Institute on Aging of the National Institutes of Health under Award Numbers R01AG081955, R21 AG073967, P30 AG062677, U01 AG006786, and R01 AG034676 (the Rochester Epidemiology Project). This work was also supported by the Kevin Merszei Career Development Award in Neurodegenerative Diseases Research IHO Janet Vittone, MD, the GHR Foundation, and the Mayo Foundation for Education and Research. The content is solely the responsibility of the authors and does not necessarily represent the official views of the National Institutes of Health or other sponsors. A Mayo Clinic invention disclosure has been submitted for the Stricker Learning Span and the Mayo Test Drive platform (NHS, JLS). We have no other conflicts of interest to disclose related to this work. The authors wish to thank the participants and staff at the Mayo Clinic Study of Aging and Mayo Alzheimer’s Disease Research Center.

## Supporting information

Supplemental File MTD Reliability

## Data Availability

All data produced in the present study are available upon reasonable request to the authors.

## Abbreviations

MTD: Mayo Test Drive
MTD: _Comp_: Mayo Test Drive Test Battery Composite Score
MTD: Stricker Learning Span
SLS_Sum_: Stricker Learning Span Sum of Trials
SYM: Symbols Test
SYM_RT_: Symbols Test Average Correct Item Response Time in Seconds
SYM_AW_: Accuracy-Weighted Score that Weights SYM_RT_ by Accuracy
CU: Cognitively Unimpaired
Mayo-PACC: Mayo Preclinical Alzheimer’s disease Cognitive Composite
AVLT: Rey’s Auditory Verbal Learning Test

## CRediT

Morgan A. Hughes: Writing – original draft, Writing – review and editing, Data curation, Visualization, Investigation

Ryan D. Frank: Conceptualization, Supervision, Data curation, Methodology, Formal analysis, Visualization

Rita L. Taylor: Supervision, Writing – review and editing

Winnie Z. Fan: Formal analysis, Visualization

Teresa J. Christianson: Data curation

Walter K. Kremers: Conceptualization, Supervision, Methodology

John L. Stricker: Data curation, Software, Writing – review and editing

Mary M. Machulda: Supervision, Writing – review and editing, Investigation

Jason Hassenstab: Conceptualization, Writing – review and editing

Michelle M. Mielke: Funding acquisition, Writing – review and editing, Supervision

John A. Lucas: Resources, Supervision, Writing – review and editing

Paula A. Aduen: Resources, Supervision, Writing – review and editing

Gregory S. Day: Resources, Writing – review and editing

Neill R. Graff-Radford: Resources, Funding acquisition, Writing – review and editing

Clifford R. Jack, Jr.: Conceptualization, Resources, Funding acquisition, Writing – review and editing

Jonathan Graff-Radford: Resources, Funding acquisition, Writing – review and editing

Ronald C. Petersen: Resources, Funding acquisition, Writing – review and editing, Supervision

Nikki H. Stricker: Conceptualization, Funding acquisition, Project administration, Methodology, Data curation, Supervision, Writing – original draft, Writing – review and editing

