## Supplemental File MTD Reliability for "Reliability of remote self-administered web-based digital cognitive measures and comparison to in-person neuropsychological tests: Stricker Learning Span, Symbols Test and the Mayo Test Drive Screening Battery Composite"

Copyright 2025 Mayo Foundation for Medical Education and Research. All materials in the Supplementary Material used with permission of Mayo Foundation of Medical Education and Research, all rights reserved.

**Supplemental Methods.**

*Change in study schedule.*

The Mayo Clinic Study of Aging shifted the study schedule during the data period presented in this study. For individuals aged 50+, the study schedule changed from visits every 15 months to visits every 18 months. This change was implemented in phases beginning in May 2024 and the phased roll-out was completed by January 1, 2025. For individuals 30-49, the study schedule changed from visits every 30 months to visits every 36 months as of January 1, 2025. To accommodate this change, the MTD session schedule changed from every 7.5 months to every 9 months as of 12/23/2024.

Mayo Alzheimer's Disease Research Center participants are seen annually for in-person visits. Remote Mayo Test Drive sessions also occur annually, shortly following the in person visit, as described in Patel et al. (2025).

For the analyses comparing MTD to in-person-administered cognitive measures, we required the last test session (the second in-person-administered cognitive session and the third MTD session) to be within 24 months of the baseline session to avoid including a session from a subsequent parent study cycle in these analyses.

**Supplemental Table 1.** Mayo Test Drive Intraclass Correlation Coefficients (ICC, 95% Confidence Interval) adjusted for age, sex, and education for all participants across all sessions and session pairs.

| MTD Measure | All 4 sessions (N=1846) | Sessions 1-2 (N=1846) | Sessions 2-3 (N=1315) | Sessions 3-4 (N=1069) |
|---|---|---|---|---|
| **Primary Variables**[1] | | | | |
| MTD Composite, raw[2] | 0.72 (0.70, 0.74) | 0.69 (0.67, 0.72) | 0.76 (0.73, 0.80) | 0.74 (0.68, 0.79) |
| SLS Sum of Trials[3] | 0.70 (0.69, 0.72) | 0.68 (0.66, 0.71) | 0.75 (0.71, 0.78) | 0.71 (0.66, 0.77) |
| SYM Accuracy-Weighted Score[4] | 0.58 (0.54, 0.61) | 0.53 (0.49, 0.58) | 0.57 (0.48, 0.66) | 0.60 (0.50, 0.69) |
| SYM Correct Items Resp. Time[5] | 0.75 (0.72, 0.77) | 0.75 (0.72, 0.78) | 0.78 (0.73, 0.83) | 0.73 (0.66, 0.80) |
| **Secondary Variables**[6] | | | | |
| MTD Composite, z-score[7] | 0.75 (0.73, 0.76) | 0.73 (0.71, 0.75) | 0.78 (0.75, 0.82) | 0.75 (0.70, 0.80) |
| SLS Max Learning Span[8] | 0.66 (0.64, 0.68) | 0.64 (0.61, 0.66) | 0.70 (0.66, 0.74) | 0.64 (0.57, 0.70) |
| SLS 1-5 Total Correct[9] | 0.67 (0.65, 0.70) | 0.65 (0.62, 0.68) | 0.73 (0.69, 0.77) | 0.68 (0.63, 0.74) |
| SLS Delay Correct | 0.67 (0.65, 0.69) | 0.66 (0.64, 0.69) | 0.67 (0.63, 0.71) | 0.68 (0.63, 0.74) |
| SYM Middle 2 Trials Avg sec.[10] | 0.74 (0.71, 0.77) | 0.74 (0.71, 0.77) | 0.77 (0.71, 0.82) | 0.71 (0.65, 0.77) |
| SYM All 4 Trials Avg sec.[11] | 0.74 (0.72, 0.77) | 0.75 (0.72, 0.78) | 0.78 (0.73, 0.83) | 0.73 (0.66, 0.80) |
| **Secondary Process Variables**[12] | | | | |
| SLS Trial 1 Correct | 0.27 (0.24, 0.31) | 0.25 (0.21, 0.30) | 0.31 (0.24, 0.38) | 0.30 (0.22, 0.39) |
| SLS Trial 2 Correct | 0.41 (0.38, 0.44) | 0.38 (0.33, 0.42) | 0.46 (0.39, 0.53) | 0.45 (0.37, 0.53) |
| SLS Trial 3 Correct | 0.54 (0.51, 0.56) | 0.52 (0.48, 0.55) | 0.57 (0.52, 0.62) | 0.52 (0.45, 0.60) |
| SLS Trial 4 Correct | 0.62 (0.60, 0.64) | 0.59 (0.56, 0.62) | 0.65 (0.60, 0.69) | 0.65 (0.60, 0.71) |
| SLS Trial 5 Correct | 0.65 (0.63, 0.67) | 0.63 (0.60, 0.65) | 0.67 (0.63, 0.71) | 0.61 (0.54, 0.67) |

| | | | | |
|---|---|---|---|---|
| SLS Percent Retention[13] | 0.16 (0.13, 0.19) | 0.17 (0.12, 0.23) | 0.11 (0.03, 0.18) | 0.19 (0.10, 0.27) |
| SYM Total Correct (Accuracy)[14] | 0.35 (0.30, 0.41) | 0.30 (0.23, 0.37) | 0.31 (0.18, 0.43) | 0.44 (0.29, 0.59) |
| SYM Trial 1 sec. | 0.59 (0.55, 0.63) | 0.58 (0.53, 0.63) | 0.64 (0.57, 0.71) | 0.63 (0.53, 0.73) |
| SYM Trial 2 sec. | 0.64 (0.61, 0.67) | 0.62 (0.58, 0.66) | 0.64 (0.57, 0.71) | 0.61 (0.53, 0.70) |
| SYM Trial 3 sec. | 0.65 (0.62, 0.68) | 0.66 (0.63, 0.70) | 0.67 (0.61, 0.73) | 0.61 (0.53, 0.68) |
| SYM Trial 4 sec. | 0.64 (0.62, 0.67) | 0.64 (0.61, 0.67) | 0.69 (0.64, 0.75) | 0.66 (0.58, 0.74) |

[1] Primary Variables are recommended for use as primary outcome variables for research studies and for clinical interpretation. The choice of which SYM outcome variable to use may differ depending on sample or individual characteristics.
[2] MTD Composite Raw = SLS Sum of Trials + SYM Accuracy-Weighted Average Response Time Correct Items
[3] SLS Sum of Trials = SLS Trials 1-5 Total Correct + SLS Delay Correct
[4] SYM Average Response Time Correct Items, Accuracy-Weighted = Symbols Average Response Time for Correct Items only weighted by accuracy (see Boots et al. [3] for details)
[5] SYM Correct Items Resp. Time = Symbols Average Response Time for Correct Items only (correct items across all 4 Symbols trials), seconds
[6] Secondary Variables can be considered for use as outcome variables for research studies and for clinical interpretation as needed (e.g., when immediate learning and delayed memory performance differentiation is needed, if session interruptions complicate interpretation of $SYM_{RT}$ or average seconds per trial).
[7] MTD composite z = [SLS Max Learning Span z + SLS 1-5 Total Correct z + SLS Delay Correct z + (-1 * SYM Correct Items Response Time z)] / 4
[8] SLS Max Learning Span = maximum number of words recognized across any of the 5 learning trials
[9] SLS 1-5 Total Correct = total words correctly recognized across all 5 learning trials
[10] SYM Middle Trials Avg Completion Time = average seconds to complete a trial when averaged across “the middle” 2 trials, excluding highest and lowest performances
[11] SYM Avg Trial Completion Time = average seconds to complete a SYM trial [(SYM Trial 1 seconds + SYM Trial 2 seconds + SYM Trial 3 seconds + SYM Trial 4 seconds) / 4].
[12] Secondary Process Variables should not be used as outcome variables for research studies. These process variables are to supplement clinical interpretation to help clinicians understand patterns of performance or to consider the potential influence of session interruptions.
[13] SLS Percent Retention = (SLS Delay Correct / SLS Max Learning Span) * 100.

[14] SYM Total Correct = total correct Symbols matches across all 4 SYM trials (accuracy); this variable is not normally distributed (most participants have high accuracy). Variables with skewed distributions typically have lower reliability.

*Note.* Avg = Average; MTD = Mayo Test Drive; Resp. Time = Response Time; SLS = Stricker Learning Span; SYM = Symbols. Winsorized raw scores were used to derive the adjusted ICCs in this table; see manuscript Table 2 for unadjusted ICCs. Table used with permission of Mayo Foundation for Medical Education and Research, all rights reserved.

**Supplemental Table 2.** Mayo Test Drive Pearson Correlation Coefficients for Winsorized raw scores (95% Confidence Interval) for all participants across session pairs.

| MTD Measure | Sessions 1-2 (N=1846) | Sessions 2-3 (N=1315) | Sessions 3-4 (N=1069) |
|---|---|---|---|
| **Primary Variables** | | | |
| MTD Composite, raw | 0.77 (0.75, 0.79) | 0.81 (0.8, 0.83) | 0.81 (0.78, 0.83) |
| SLS Sum of Trials | 0.74 (0.72, 0.76) | 0.78 (0.75, 0.8) | 0.77 (0.74, 0.79) |
| SYM Accuracy-Weighted Score | 0.66 (0.64, 0.69) | 0.72 (0.69, 0.74) | 0.75 (0.72, 0.77) |
| SYM Correct Items Resp. Time | 0.83 (0.82, 0.84) | 0.85 (0.83, 0.86) | 0.85 (0.84, 0.87) |
| **Secondary Variables** | | | |
| MTD Composite, z-score | 0.80 (0.78, 0.81) | 0.83 (0.81, 0.84) | 0.82 (0.8, 0.84) |
| SLS Max Learning Span | 0.69 (0.67, 0.71) | 0.73 (0.7, 0.75) | 0.71 (0.68, 0.74) |
| SLS 1-5 Total Correct | 0.71 (0.69, 0.73) | 0.76 (0.73, 0.78) | 0.74 (0.71, 0.76) |
| SLS Delay Correct | 0.71 (0.69, 0.73) | 0.73 (0.7, 0.75) | 0.73 (0.71, 0.76) |
| SYM Middle 2 Trials Avg sec. | 0.82 (0.81, 0.84) | 0.84 (0.82, 0.85) | 0.84 (0.82, 0.86) |
| SYM All 4 Trials Avg sec. | 0.83 (0.81, 0.84) | 0.85 (0.83, 0.86) | 0.85 (0.83, 0.87) |
| **Secondary Process Variables** | | | |
| SLS Trial 1 Correct | 0.32 (0.28, 0.36) | 0.36 (0.31, 0.41) | 0.33 (0.28, 0.39) |
| SLS Trial 2 Correct | 0.46 (0.43, 0.50) | 0.52 (0.48, 0.55) | 0.51 (0.46, 0.55) |
| SLS Trial 3 Correct | 0.58 (0.55, 0.61) | 0.62 (0.59, 0.65) | 0.60 (0.56, 0.64) |
| SLS Trial 4 Correct | 0.65 (0.62, 0.67) | 0.69 (0.66, 0.72) | 0.71 (0.68, 0.74) |
| SLS Trial 5 Correct | 0.68 (0.65, 0.70) | 0.70 (0.68, 0.73) | 0.68 (0.65, 0.72) |
| SLS Percent Retention | 0.19 (0.14, 0.23) | 0.16 (0.10, 0.21) | 0.21 (0.16, 0.27) |
| SYM Total Correct (Accuracy) | 0.32 (0.28, 0.36) | 0.40 (0.35, 0.44) | 0.46 (0.41, 0.50) |
| SYM Trial 1 sec. | 0.69 (0.66, 0.71) | 0.73 (0.70, 0.75) | 0.76 (0.73, 0.78) |
| SYM Trial 2 sec. | 0.72 (0.70, 0.74) | 0.73 (0.71, 0.76) | 0.77 (0.75, 0.80) |
| SYM Trial 3 sec. | 0.76 (0.74, 0.78) | 0.77 (0.75, 0.79) | 0.74 (0.71, 0.77) |
| SYM Trial 4 sec. | 0.74 (0.72, 0.76) | 0.77 (0.75, 0.79) | 0.77 (0.75, 0.80) |

*Note*. See manuscript Table 2 or Supplemental Table 1 for variable definitions. Pearson correlation coefficients are provided because these are frequently used as reliability coefficients in neuropsychology and are easily reproducible (whereas there are numerous methods of calculating ICC values). Table used with permission of Mayo Foundation for Medical Education and Research, all rights reserved.

**Supplemental Table 3.** Mayo Test Drive Pearson Correlation Coefficients for non-Winsorized raw scores (95% Confidence Interval) for all participants across session pairs.

| MTD Measure | Sessions 1-2 (N=1846) | Sessions 2-3 (N=1315) | Sessions 3-4 (N=1069) |
|---|---|---|---|
| **Primary Variables** | | | |
| MTD Composite, raw | 0.77 (0.76, 0.79) | 0.82 (0.8, 0.83) | 0.81 (0.78, 0.83) |
| SLS Sum of Trials | 0.74 (0.72, 0.76) | 0.78 (0.75, 0.80) | 0.77 (0.74, 0.79) |
| SYM Accuracy-Weighted Score | 0.67 (0.64, 0.69) | 0.72 (0.70, 0.75) | 0.75 (0.72, 0.78) |
| SYM Correct Items Resp. Time | 0.81 (0.79, 0.82) | 0.81 (0.79, 0.83) | 0.83 (0.81, 0.85) |
| **Secondary Variables** | | | |
| MTD Composite, z-score | 0.80 (0.78, 0.82) | 0.83 (0.81, 0.84) | 0.82 (0.80, 0.84) |
| SLS Max Learning Span | 0.69 (0.66, 0.71) | 0.73 (0.70, 0.75) | 0.71 (0.68, 0.74) |
| SLS 1-5 Total Correct | 0.71 (0.69, 0.73) | 0.76 (0.73, 0.78) | 0.74 (0.71, 0.76) |
| SLS Delay Correct | 0.71 (0.69, 0.73) | 0.73 (0.70, 0.75) | 0.74 (0.71, 0.76) |
| SYM Middle 2 Trials Avg sec. | 0.80 (0.78, 0.82) | 0.81 (0.79, 0.83) | 0.82 (0.79, 0.83) |
| SYM All 4 Trials Avg sec. | 0.81 (0.79, 0.82) | 0.81 (0.79, 0.82) | 0.79 (0.76, 0.81) |
| **Secondary Process Variables** | | | |
| SLS Trial 1 Correct | 0.32 (0.28, 0.36) | 0.36 (0.31, 0.41) | 0.34 (0.28, 0.39) |
| SLS Trial 2 Correct | 0.46 (0.43, 0.50) | 0.52 (0.48, 0.56) | 0.51 (0.46, 0.55) |
| SLS Trial 3 Correct | 0.58 (0.55, 0.61) | 0.62 (0.59, 0.65) | 0.60 (0.56, 0.64) |
| SLS Trial 4 Correct | 0.65 (0.62, 0.67) | 0.69 (0.66, 0.72) | 0.71 (0.68, 0.74) |
| SLS Trial 5 Correct | 0.68 (0.65, 0.70) | 0.71 (0.68, 0.73) | 0.69 (0.65, 0.72) |
| SLS Percent Retention | 0.19 (0.14, 0.23) | 0.15 (0.10, 0.21) | 0.22 (0.16, 0.28) |
| SYM Total Correct (Accuracy) | 0.31 (0.27, 0.35) | 0.40 (0.36, 0.45) | 0.48 (0.44, 0.53) |
| SYM Trial 1 sec. | 0.66 (0.63, 0.68) | 0.68 (0.65, 0.71) | 0.57 (0.53, 0.61) |
| SYM Trial 2 sec. | 0.70 (0.68, 0.73) | 0.66 (0.63, 0.69) | 0.75 (0.72, 0.77) |
| SYM Trial 3 sec. | 0.66 (0.64, 0.69) | 0.72 (0.69, 0.75) | 0.71 (0.68, 0.74) |
| SYM Trial 4 sec. | 0.72 (0.70, 0.74) | 0.75 (0.73, 0.77) | 0.78 (0.76, 0.80) |

*Note*. See manuscript Table 2 or Supplemental Table 1 for variable definitions. Table used with permission of Mayo Foundation for Medical Education and Research, all rights reserved.

**Supplemental Table 4**. Intraclass Correlation Coefficients (ICC, 95% Confidence Interval) comparisons of in-person-administered traditional neuropsychological measures versus remote self-administered MTD reliability values for a subset of 244 participants who completed 2 in-person tests and 3 remote tests.

| | Unadjusted | | | Adjusted | | |
|---|---|---|---|---|---|---|
| **Outcome** | **In-person ICC (95% CI)** | **Remote ICC (95% CI)** | **p** | **In-person ICC (95% CI)** | **Remote ICC (95% CI)** | **p** |
| **Sessions 1-2 vs 1-2** | | | | | | |
| STMS vs MTD composite | 0.67 (0.58, 0.75) | 0.75 (0.70, 0.81) | 0.06 | 0.57 (0.47, 0.66) | 0.66 (0.59, 0.73) | 0.10 |
| Mayo-PACC vs MTD composite | 0.80 (0.75, 0.85) | 0.75 (0.70, 0.81) | 0.25 | 0.65 (0.57, 0.73) | 0.66 (0.59, 0.73) | 0.89 |
| AVLT vs SLS sum of trials | 0.78 (0.73, 0.83) | 0.72 (0.66, 0.79) | 0.15 | 0.64 (0.57, 0.72) | 0.65 (0.58, 0.73) | 0.81 |
| Trails B vs $SYM_{AW}$ | 0.74 (0.66, 0.82) | 0.61 (0.50, 0.71) | 0.041 | 0.61 (0.51, 0.72) | 0.47 (0.34, 0.60) | 0.10 |
| Trails B vs $SYM_{RT}$ | 0.74 (0.66, 0.82) | 0.83 (0.78, 0.88) | 0.051 | 0.61 (0.51, 0.72) | 0.75 (0.68, 0.82) | 0.025 |
| **Sessions 1-2 vs 1-3** | | | | | | |
| STMS vs MTD Composite | 0.67 (0.58, 0.75) | 0.76 (0.70, 0.81) | 0.07 | 0.57 (0.47, 0.66) | 0.67 (0.60, 0.74) | 0.09 |
| Mayo-PACC vs MTD Composite | 0.80 (0.75, 0.85) | 0.76 (0.70, 0.81) | 0.18 | 0.65 (0.57, 0.73) | 0.67 (0.60, 0.74) | 0.72 |
| AVLT vs SLS Sum of Trials | 0.78 (0.73, 0.83) | 0.73 (0.67, 0.79) | 0.17 | 0.64 (0.57, 0.72) | 0.67 (0.61, 0.74) | 0.49 |
| Trails B vs $SYM_{AW}$ | 0.74 (0.66, 0.82) | 0.60 (0.48, 0.73) | 0.09 | 0.61 (0.51, 0.72) | 0.46 (0.31, 0.61) | 0.12 |
| Trails B vs $SYM_{RT}$ | 0.74 (0.66, 0.82) | 0.79 (0.73, 0.86) | 0.24 | 0.61 (0.51, 0.72) | 0.68 (0.59, 0.77) | 0.29 |
| **Sessions 1-2 vs 2-3** | | | | | | |
| STMS vs MTD Composite | 0.67 (0.58, 0.75) | 0.78 (0.73, 0.83) | 0.026 | 0.57 (0.47, 0.66) | 0.70 (0.63, 0.76) | 0.040 |
| Mayo-PACC vs MTD Composite | 0.80 (0.75, 0.85) | 0.78 (0.73, 0.83) | 0.51 | 0.65 (0.57, 0.73) | 0.70 (0.63, 0.76) | 0.39 |
| AVLT vs SLS Sum of Trials | 0.78 (0.73, 0.83) | 0.76 (0.70, 0.82) | 0.52 | 0.64 (0.57, 0.72) | 0.70 (0.63, 0.77) | 0.18 |
| Trails B vs $SYM_{AW}$ | 0.74 (0.66, 0.82) | 0.59 (0.47, 0.71) | 0.06 | 0.61 (0.51, 0.72) | 0.44 (0.29, 0.59) | 0.09 |

| | | | | | | |
|---|---|---|---|---|---|---|
| Trails B vs $SYM_{RT}$ | 0.74 (0.66, 0.82) | 0.78 (0.72, 0.84) | 0.41 | 0.61 (0.51, 0.72) | 0.66 (0.56, 0.75) | 0.49 |
| **Sessions 1-2 vs 1-2-3** | | | | | | |
| STMS vs MTD Composite | 0.67 (0.58, 0.75) | 0.76 (0.72, 0.81) | 0.034 | 0.57 (0.47, 0.66) | 0.68 (0.62, 0.73) | 0.050 |
| Mayo-PACC vs MTD Composite | 0.80 (0.75, 0.85) | 0.76 (0.72, 0.81) | 0.24 | 0.65 (0.57, 0.73) | 0.68 (0.62, 0.73) | 0.60 |
| AVLT vs SLS Sum of Trials | 0.78 (0.73, 0.83) | 0.74 (0.69, 0.79) | 0.19 | 0.64 (0.57, 0.72) | 0.68 (0.62, 0.73) | 0.39 |
| Trails B vs $SYM_{AW}$ | 0.74 (0.66, 0.82) | 0.60 (0.50, 0.70) | 0.035 | 0.61 (0.51, 0.72) | 0.46 (0.35, 0.57) | 0.07 |
| Trails B vs $SYM_{RT}$ | 0.74 (0.66, 0.82) | 0.80 (0.75, 0.85) | 0.16 | 0.61 (0.51, 0.72) | 0.70 (0.63, 0.76) | 0.14 |

*Note*. AVLT Sum of Trials = Rey's Auditory Verbal Learning Test trials 1-5 correct + trial 6 correct + 30-minute delayed recall correct. Mayo-PACC = Mayo Preclinical Alzheimer's disease Cognitive Composite (average z of Auditory Verbal Learning Test sum of trials, animal fluency, and inversed Trails B). MTD Composite = MTD raw composite (SLS Sum of Trials + $SYM_{AW}$). STMS = Kokmen Short Test of Mental Status (a multi-domain screening measure similar to the MMSE). $SYM_{AW}$ = Symbols Accuracy-Weighted Score. $SYM_{RT}$ = Symbols average correct item response time. Winsorized raw scores were used to derive the unadjusted ICCs in this table. This subsample of individuals test naïve at their in-person baseline included participants 71.3 (SD=10.3) years old with 15.8 (SD=2.3) years of education, 45.5% were male, 98.3% were White, 100% were non-Hispanic, and 97.5% were cognitively unimpaired. Table used with permission of Mayo Foundation for Medical Education and Research, all rights reserved.

**Supplemental Table 5.** Mayo Test Drive Intraclass Correlation Coefficients (ICC, 95% Confidence Interval) for analyses comparing subgroups of Cognitively Unimpaired participants to explore the impact of demographic variables and factors relevant for remote self-administered digital cognitive assessment.

| | **Unadjusted ICCs** | | | **Adjusted ICCs (adjust for age, sex and educ)** | | |
|---|---|---|---|---|---|---|
| | **Group 1** | **Group 2** | | **Group 1** | **Group 2** | |
| **Outcome** | **ICC (95% CI)** | **ICC (95% CI)** | **p** | **ICC (95% CI)** | **ICC (95% CI)** | **p** |
| **By Sex** | Female (N=940) | Male (N=846) | - | Female (N=940) | Male (N=846) | - |
| MTD Composite, raw | 0.75 (0.73, 0.78) | 0.76 (0.73, 0.79) | 0.66 | 0.70 (0.67, 0.73) | 0.70 (0.67, 0.74) | 0.91 |
| SLS Sum of Trials | 0.72 (0.69, 0.75) | 0.73 (0.70, 0.75) | 0.73 | 0.69 (0.65, 0.72) | 0.69 (0.65, 0.72) | 0.98 |
| SYM Accuracy-Weighted Score | 0.66 (0.62, 0.70) | 0.69 (0.64, 0.74) | 0.41 | 0.53 (0.47, 0.59) | 0.57 (0.51, 0.64) | 0.30 |
| SYM Correct Items Resp. Time | 0.83 (0.80, 0.85) | 0.81 (0.78, 0.84) | 0.37 | 0.74 (0.70, 0.78) | 0.72 (0.68, 0.76) | 0.47 |
| **By Education** | <Bachelors (N=685) | Bachelors+ (N=1099) | - | <Bachelors (N=685) | Bachelors+ (N=1099) | - |
| MTD Composite, raw | 0.76 (0.73, 0.79) | 0.77 (0.74, 0.79) | 0.65 | 0.69 (0.65, 0.73) | 0.70 (0.68, 0.73) | 0.67 |
| SLS Sum of Trials | 0.72 (0.69, 0.76) | 0.73 (0.71, 0.76) | 0.57 | 0.68 (0.64, 0.72) | 0.70 (0.67, 0.72) | 0.45 |
| SYM Accuracy-Weighted Score | 0.64 (0.59, 0.70) | 0.70 (0.66, 0.74) | 0.11 | 0.54 (0.47, 0.60) | 0.57 (0.52, 0.62) | 0.40 |
| SYM Correct Items Resp. Time | 0.79 (0.76, 0.83) | 0.83 (0.81, 0.85) | 0.07 | 0.72 (0.67, 0.76) | 0.74 (0.71, 0.77) | 0.35 |
| **By Age** | <70 at first (N=898) | 70+ at first (N=888) | - | <70 at first (N=898) | 70+ at first (N=888) | - |
| MTD Composite, raw | 0.72 (0.69, 0.75) | 0.75 (0.73, 0.78) | 0.10 | 0.69 (0.65, 0.72) | 0.74 (0.71, 0.76) | 0.027 |
| SLS Sum of Trials | 0.71 (0.68, 0.74) | 0.72 (0.70, 0.75) | 0.49 | 0.68 (0.65, 0.71) | 0.71 (0.68, 0.74) | 0.21 |
| SYM Accuracy-Weighted Score | 0.53 (0.45, 0.60) | 0.65 (0.60, 0.69) | 0.008 | 0.51 (0.44, 0.59) | 0.64 (0.59, 0.68) | 0.007 |
| SYM Correct Items Resp. Time | 0.74 (0.71, 0.77) | 0.78 (0.75, 0.81) | 0.06 | 0.73 (0.70, 0.77) | 0.78 (0.75, 0.81) | 0.054 |
| **By Device** | Consistent (N=1310) | Inconsistent (N=476) | - | Consistent (N=1310) | Inconsistent (N=476) | - |

| | | | | | | |
|---|---|---|---|---|---|---|
| MTD Composite, raw | 0.77 (0.75, 0.79) | 0.76 (0.72, 0.79) | 0.45 | 0.71 (0.69, 0.73) | 0.67 (0.63, 0.72) | 0.13 |
| SLS Sum of Trials | 0.74 (0.72, 0.76) | 0.73 (0.69, 0.77) | 0.79 | 0.70 (0.67, 0.72) | 0.67 (0.63, 0.72) | 0.34 |
| SYM Accuracy-Weighted Score | 0.68 (0.65, 0.72) | 0.64 (0.57, 0.72) | 0.32 | 0.57 (0.53, 0.61) | 0.50 (0.40, 0.59) | 0.16 |
| SYM Correct Items Resp. Time | 0.83 (0.80, 0.85) | 0.79 (0.75, 0.83) | 0.10 | 0.74 (0.71, 0.77) | 0.68 (0.62, 0.74) | 0.049 |
| **Any Interference** | No (N=1189) | Yes (N=597) | - | No (N=1189) | Yes (N=597) | - |
| MTD Composite, raw | 0.76 (0.74, 0.78) | 0.77 (0.74, 0.80) | 0.63 | 0.69 (0.67, 0.72) | 0.71 (0.67, 0.75) | 0.49 |
| SLS Sum of Trials | 0.73 (0.71, 0.76) | 0.74 (0.70, 0.77) | 0.93 | 0.69 (0.66, 0.71) | 0.69 (0.65, 0.73) | 0.78 |
| SYM Accuracy-Weighted Score | 0.69 (0.65, 0.72) | 0.66 (0.60, 0.72) | 0.38 | 0.56 (0.52, 0.61) | 0.52 (0.46, 0.59) | 0.28 |
| SYM Correct Items Resp. Time | 0.83 (0.81, 0.85) | 0.80 (0.76, 0.84) | 0.13 | 0.75 (0.72, 0.77) | 0.69 (0.63, 0.75) | 0.10 |
| **By Location** | All home (N=1516) | All not home (N=270) | - | All home (N=1516) | All not home (N=270) | - |
| MTD Composite, raw | 0.77 (0.75, 0.79) | 0.71 (0.65, 0.77) | 0.06 | 0.71 (0.68, 0.73) | 0.65 (0.59, 0.72) | 0.14 |
| SLS Sum of Trials | 0.74 (0.72, 0.76) | 0.68 (0.63, 0.74) | 0.08 | 0.69 (0.67, 0.72) | 0.65 (0.58, 0.71) | 0.17 |
| SYM Accuracy-Weighted Score | 0.68 (0.65, 0.71) | 0.58 (0.50, 0.66) | 0.025 | 0.56 (0.52, 0.61) | 0.45 (0.38, 0.53) | 0.009 |
| SYM Correct Items Resp. Time | 0.82 (0.80, 0.84) | 0.76 (0.71, 0.81) | 0.048 | 0.74 (0.71, 0.76) | 0.65 (0.59, 0.71) | 0.006 |

*Note*. Table used with permission of Mayo Foundation for Medical Education and Research, all rights reserved. See manuscript for details about how subgroups were defined. Descriptives for subgroups are provided in Supplementary Tables 6 and 7. Significant differences by group were as follows: <70-year-olds had significantly higher mean years of education compared to 70+ (15.9 vs 15.4, p<.001). Males had significantly higher years of mean education vs females (16.0 vs 15.4, p<.001). Those with 16+ yrs of education had significantly lower mean age (68.5 vs 71.6, p<.001) and were more likely male (53.3% vs 39.3%, p<.001) than those with <16 yrs of education. Inconsistent device users were significantly younger (68.2 vs 70.3, p<.001) compared to consistent users. Those reporting interference had lower mean age (68.9 vs 70.2, p=0.03) and were more likely female (58.0% vs 49.2%, <.001) compared to those with interference. Those who reported taking all their MTDs at home were significantly older (71.5 vs 59.7, p<.001) and had fewer years of education (15.6 vs 16.2, p<.001) vs those not all at home.

**Supplemental Table 6**. Age, Sex, Education descriptive statistics by demographic categories for analyses comparing subgroups of Cognitively Unimpaired participants (n=1786) to explore the impact of demographic variables and factors relevant for remote self-administered digital cognitive assessment.

| | By Age | | | By Sex | | | By Education | | |
|---|---|---|---|---|---|---|---|---|---|
| Characteristic | <70 N=898 | 70+ N=888 | p-value | Female N=940 | Male N=846 | p-value | <Bachelors N=685 | >=Bachelors N=1099 | p-value |
| **Age at MTD** | 60.1 (8.4) | 79.2 (6.2) | <0.001 † | 69.6 (12.0) | 69.8 (12.1) | 0.76 † | 71.6 (11.6) | 68.5 (12.2) | <0.001 † |
| **Sex** | | | 0.94 ‡ | | | <0.001 ‡ | | | <0.001 ‡ |
| Female | 475 (52.1%) | 488 (52.2%) | | 963 (100.0%) | 0 (0.0%) | - | 436 (60.7%) | 526 (46.7%) | |
| Male | 437 (47.9%) | 446 (47.8%) | | 0 (0.0%) | 883 (100.0%) | | 282 (39.3%) | 600 (53.3%) | |
| **Education** | 15.9 (2.2) | 15.4 (2.4) | <0.001 † | 15.4 (2.3) | 16.0 (2.3) | <0.001 † | 13.2 (1.1) | 17.2 (1.3) | <0.001 † |

*Note*. Table used with permission of Mayo Foundation for Medical Education and Research, all rights reserved.

**Supplemental Table 7**. Age, Sex, Education descriptive statistics by demographic categories for analyses comparing subgroups of Cognitively Unimpaired participants (n=1844) to explore the impact of factors relevant for remote self-administered digital cognitive assessment.

| | By Device | | | By Interference | | | By Location | | |
|---|---|---|---|---|---|---|---|---|---|
| Characteristic | Inconsistent N=476 | Consistent N=1310 | p-value | No N=1189 | Yes N=597 | p-value | Not All Home N=270 | All Home N=1516 | p-value |
| **Age at MTD** | 68.2 (12.0) | 70.3 (12.0) | <0.001 † | 70.2 (11.9) | 68.9 (12.3) | 0.03 † | 59.7 (12.0) | 71.5 (11.2) | <0.001 † |
| **Sex** | | | 0.81 ‡ | | | <0.001 ‡ | | | 0.61 ‡ |
| Female | 256 (51.7%) | 707 (52.3%) | | 605 (49.2%) | 358 (58.0%) | | 138 (50.7%) | 825 (52.4%) | |
| Male | 239 (48.3%) | 644 (47.7%) | | 624 (50.8%) | 259 (42.0%) | | 134 (49.3%) | 749 (47.6%) | |
| **Education** | 15.8 (2.3) | 15.6 (2.3) | 0.18 † | 15.6 (2.3) | 15.7 (2.3) | 0.72 † | 16.2 (2.5) | 15.6 (2.3) | <0.001 † |

*Note*. Table used with permission of Mayo Foundation for Medical Education and Research, all rights reserved.
